# A Systematic Review of the Application of Computational Grounded Theory Method in Healthcare Research

**DOI:** 10.1101/2025.07.02.25330783

**Authors:** Ravi Shankar, Fiona Devi, Xu Qian

## Abstract

The integration of computational methods with traditional qualitative research approaches has emerged as a transformative paradigm in healthcare research. Computational Grounded Theory (CGT) represents a novel methodological framework that combines the interpretive depth of grounded theory with the analytical power of computational techniques, particularly machine learning and natural language processing. This systematic review examines the application of CGT in healthcare research, synthesizing evidence from eight key studies that demonstrate the method’s utility across diverse healthcare contexts.

The primary objectives of this review are to analyze the current state of CGT application in healthcare research, evaluate the methodological approaches and computational techniques employed, identify key themes and patterns emerging from CGT studies, assess the advantages and limitations compared to traditional qualitative methods, and provide recommendations for future research and practice. Following a systematic search and narrative synthesis approach, eight papers applying CGT methods in healthcare contexts were analyzed, examining methodological frameworks, computational techniques, healthcare applications, and outcomes.

The results demonstrate diverse applications of CGT across healthcare domains including COVID-19 risk perception, medical AI adoption, mental health interventions, diabetes management, women’s health technology, online health communities, and old-age social welfare systems. Common computational techniques included Latent Dirichlet Allocation (LDA), sentiment analysis, word embeddings, and various machine learning algorithms. Studies consistently reported enhanced analytical capacity, identification of latent patterns, and novel theoretical insights as compared to traditional methods. However, challenges included technical complexity, interpretation validity, and the need for interdisciplinary expertise.

This review concludes that CGT represents a promising methodological innovation for healthcare research, offering unique capabilities for analyzing large-scale textual data while maintaining theoretical depth. The approach demonstrates particular value in understanding complex healthcare phenomena, patient experiences, and technology adoption. Future research should focus on standardizing methodological procedures, developing best practices, and expanding applications to emerging healthcare challenges.

## Introduction

Healthcare research increasingly confronts the challenge of analyzing vast amounts of unstructured textual data generated through electronic health records, social media platforms, patient forums, and digital health applications. Traditional qualitative research methods, while providing rich interpretive insights, often struggle with the scale and complexity of contemporary healthcare data. Simultaneously, purely computational approaches may lack the theoretical depth and contextual understanding essential for meaningful healthcare insights (Nelson, 2020). This methodological gap has become particularly evident as healthcare systems globally embrace digital transformation, generating unprecedented volumes of textual data that contain valuable insights into health behaviors, treatment experiences, and system dynamics.

Computational Grounded Theory (CGT) emerges as a methodological bridge, integrating the interpretive rigor of grounded theory with computational techniques from machine learning and natural language processing. First articulated by Nelson (2020), CGT represents a systematic approach to pattern detection, interpretive analysis, and theory development that leverages both human expertise and computational power. The framework addresses a fundamental challenge in contemporary research: how to maintain the theoretical sensitivity and contextual understanding of qualitative methods while harnessing the analytical capacity of computational approaches to process large-scale data.

The healthcare domain presents unique challenges and opportunities for CGT application. Healthcare data encompasses diverse formats including clinical narratives, patient experiences, professional communications, and health-related social media content. These data sources contain valuable insights into health behaviors, treatment experiences, professional practices, and healthcare system dynamics. However, their volume and complexity often exceed the capacity of traditional qualitative analysis methods. For instance, a single healthcare forum may generate thousands of posts daily, each containing nuanced perspectives on health conditions, treatments, and care experiences that would be impossible to analyze manually in a timely manner.

The significance of developing appropriate methodological frameworks for analyzing healthcare text data extends beyond academic considerations. As healthcare systems globally face challenges such as aging populations, rising chronic disease burdens, and rapid technological change, the ability to derive timely insights from diverse data sources becomes critical for evidence-based policy and practice. CGT offers a methodological framework that can inform real-time decision-making while maintaining the depth of understanding necessary for addressing complex healthcare challenges.

This systematic review examines eight key studies that have applied CGT methods in healthcare research contexts. These studies span diverse healthcare domains and demonstrate various computational techniques, providing a comprehensive overview of the current state of CGT application in healthcare. By synthesizing these studies, this review aims to establish the methodological foundations, identify best practices, and chart future directions for CGT in healthcare research. The review addresses critical questions about how CGT methods are being applied, what insights they generate, and how they compare to traditional approaches in addressing healthcare research questions.

## Methodology

This systematic review followed an adapted PRISMA framework suitable for methodological reviews, with modifications to accommodate the focus on methodological innovation rather than intervention effectiveness. The review process was designed to comprehensively analyze the application of CGT methods in healthcare research, examining both methodological approaches and substantive findings.

A comprehensive systematic search strategy aimed to identify all studies applying CGT methods in healthcare research contexts across multiple databases. The systematic search was conducted in May 2025 across five major electronic databases. These included PubMed/MEDLINE, Web of Science Core Collection, IEEE Xplore Digital Library, and ACM Digital Library, covering the period till 2025. Additionally, Google Scholar was searched, with the first 200 results reviewed for each search string to ensure comprehensive coverage of relevant literature.

The included studies were selected based on predetermined criteria ensuring methodological rigor and relevance to healthcare contexts. Studies were required to demonstrate application of computational methods consistent with CGT principles, including pattern detection through computational techniques and interpretive refinement by human analysts. All studies needed to focus on healthcare-related research contexts, demonstrate clear integration of pattern detection and interpretive analysis, and be published in peer-reviewed venues to ensure quality.

The search strategy employed a combination of targeted search terms and defined eligibility criteria to identify relevant studies. Search terms included variations of CGT and related methodologies within healthcare contexts, such as: (“computational grounded theory” OR “computational GT”) AND (health^*^ OR medical OR clinical OR patient^*^); (“topic modeling” OR “LDA” OR “latent dirichlet”) AND (“grounded theory”) AND (health^*^ OR medical); and (“machine learning” AND “qualitative analysis”) AND (health^*^ OR medical).

Studies were included if they applied CGT methods, were conducted in healthcare or medical research contexts, were published between till 2025 (reflecting the period when CGT methodology was established), were peer-reviewed. Exclusion criteria were studies that used only traditional grounded theory methods, focused on non-healthcare domains, were limited to conference abstracts without full papers, or were systematic reviews or meta-analyses.

The study selection process followed a systematic and transparent approach using Covidence systematic review software for both screening and data extraction. The initial database search yielded 1,247 records. After removing duplicates, 892 unique records remained. These were screened by title and abstract, resulting in 84 potentially eligible articles. Full-text assessment of these 84 articles led to the final inclusion of 8 studies that met all inclusion criteria. Two reviewers independently conducted the title and abstract screening, with any conflicts resolved through discussion. Full-text evaluations were also independently carried out by both reviewers, guided by the predetermined inclusion criteria.

A structured data extraction framework was developed to capture key methodological and substantive elements from each study. Study characteristics including authors, year of publication, healthcare domain, and primary research questions were systematically recorded. The methodological approach employed in each study was analyzed, focusing on computational techniques used, analytical procedures followed, and methods for integrating human interpretation with computational outputs. Data sources were characterized by type, volume, and characteristics of textual data analyzed. Key findings from each study were extracted, including substantive insights about healthcare phenomena, theoretical contributions to understanding healthcare issues, and methodological innovations in applying CGT. Finally, strengths and limitations of each study were assessed, particularly focusing on advantages over traditional methods and challenges encountered in implementation.

Given the methodological focus of this review, a narrative synthesis approach was employed to integrate findings across studies. This approach allowed for detailed examination of methodological variations, contextual factors, and emergent patterns in CGT application. The synthesis was organized into thematic categories addressing the review’s research objectives, enabling identification of common approaches, divergent strategies, and best practices across studies.

Quality assessment of the included studies was conducted using adapted criteria tailored for computational qualitative research. This evaluation focused on five key dimensions: methodological transparency and reproducibility; integration of computational and interpretive approaches; theoretical contribution and grounding; validity of computational procedures; and the practical applicability of findings. Each study was independently assessed and rated as high, moderate, or low quality across these dimensions to ensure a consistent and rigorous appraisal of methodological robustness and relevance.

## Results

### Overview of Included Studies

Following systematic screening, eight studies met inclusion criteria and were included in the final analysis. These studies demonstrate the breadth of CGT application across healthcare domains, methodological approaches, and geographical contexts (Table 1). Each study contributes unique insights into both the methodological development of CGT and its application to substantive healthcare challenges. Adekunle et al. (2025) explored COVID-19 risk conceptualizations in online communities using Reddit data, analyzing 500,000 comments to understand how different ideological communities construct understanding of pandemic risks. Yue et al. (2025) developed a comprehensive tutorial on using ChatGPT for grounded theory analysis in healthcare contexts, providing practical guidance for researchers seeking to integrate AI assistance into qualitative research. Guo et al. (2024) analyzed FemTech affordances through examination of period and fertility app reviews, identifying key features and user experiences that shape technology adoption.

**Table 1.** Summary of Included Studies.

| Study | Healthcare Domain | Data Source | Sample Size | Primary Computational Method | Key Innovation |
| --- | --- | --- | --- | --- | --- |
| Adekunle et al. (2025) | COVID-19 risk perception | Reddit (r/medicine) | 500,000 comments | LDA topic modeling | Ideological community analysis |
| Yue et al. (2025) | AI in qualitative research | Semi-structured interviews | 31 interviews | ChatGPT-assisted coding | Human-AI collaboration framework |
| Guo et al. (2024) | Women's health technology | App reviews | 21,489 reviews | LDA topic modeling | Affordance theory application |
| Vidyadharan et al. (2024) | Diabetes management | Mixed qualitative data | Not specified | NLP & Deep learning | Semi-supervised learning approach |
| Figueroa et al. (2023) | Health equity/racism | Medical literature | 31 articles | Word embeddings | Racism narrative framework |
| Berriche et al. (2024) | Health campaigns | Twitter | 144,906 tweets | LDA + Sentiment analysis | Engagement segmentation |
| Weber et al. (2024) | AI adoption | Reddit (medical subs) | 181 threads | Clustering + thematic analysis | Mixed-method integration |
| Agudamu et al. (2025) | Old-age welfare | Chinese literature | 413 articles | Modified LDA | Policy theme identification |

Additional studies expanded the application domains further. Vidyadharan et al. (2024) applied CGT to diabetes prevention and management using natural language processing techniques, demonstrating the potential for identifying evidence-based interventions from qualitative data. Figueroa et al. (2023) examined racism narratives in medical literature, developing a comprehensive framework for understanding how racial health inequities are discussed in academic publications. Berriche et al. (2024) studied the #Movember health campaign through computational analysis of tweets, revealing patterns of engagement and awareness. Weber et al. (2024) investigated medical professionals’ perceptions of AI using Reddit data, uncovering complex attitudes toward technology adoption. Finally, Agudamu et al. (2025) analyzed China’s old-age social welfare system through examination of academic literature, identifying key policy themes and recommendations.

### Computational Techniques Employed

The studies demonstrated diverse computational approaches within the CGT framework, with topic modeling emerging as the most commonly employed technique (Table 2). Six out of eight studies utilized Latent Dirichlet Allocation as their primary computational method for pattern detection. LDA enabled researchers to identify latent themes in large textual corpora without predetermined categories, allowing for discovery of unexpected patterns and relationships. For instance, Adekunle et al. (2025) used LDA to identify 10 topics from 500,000 Reddit comments, revealing distinct conceptualizations of COVID-19 risk across ideologically different communities. The selection of optimal topic numbers varied across studies, with researchers employing various metrics including perplexity, coherence scores, and human judgment to determine appropriate granularity.

**Table 2.** Computational Techniques and Their Applications.

| Technique | Frequency | Studies Using | Primary Purpose | Advantages | Limitations |
| --- | --- | --- | --- | --- | --- |
| Latent Dirichlet Allocation (LDA) | 6/8 studies | Adekunle, Guo, Berriche, Agudamu, Figueroa, Weber | Topic discovery | Unsupervised pattern detection | Requires topic number selection |
| Sentiment Analysis | 3/8 studies | Berriche, Weber, Figueroa | Emotional valence | Captures affective dimensions | May miss context |
| Word Embeddings | 2/8 studies | Figueroa, Vidyadharan | Semantic relationships | Context-aware representations | Computationally intensive |
| Deep Learning/NLP | 2/8 studies | Vidyadharan, Yue | Complex pattern recognition | Handles non-linear patterns | Requires large datasets |
| Clustering | 1/8 studies | Weber | Group identification | Identifies natural groupings | Sensitive to parameters |
| ChatGPT/LLM | 1/8 studies | Yue | Coding assistance | Enhances efficiency | Requires validation |

Sentiment analysis was incorporated in multiple studies to understand emotional dimensions of healthcare experiences. Berriche et al. (2024) combined LDA with sentiment analysis to examine 144,906 tweets about the #Movember campaign, identifying both collective emotions and cognitive factors influencing health behavior engagement. This combination of techniques enabled researchers to move beyond simple topic identification to understand the emotional valence associated with different health topics, providing insights into barriers and facilitators of health behavior change.

Advanced natural language processing techniques were employed to address specific analytical challenges. Vidyadharan et al. (2024) utilized deep learning approaches for analyzing qualitative data about diabetes management, demonstrating the potential for semi-supervised learning in healthcare contexts where labeled data may be limited. Their approach combined traditional NLP techniques with neural network architectures to identify complex patterns in patient narratives about diabetes self-management. Weber et al. (2024) exemplified sophisticated integration of multiple computational techniques, combining unsupervised clustering with qualitative thematic analysis to understand medical professionals’ AI perceptions. This multi-method approach enabled identification of nuanced attitudes that might be missed by single-technique analyses.

Several studies employed novel computational approaches tailored to their specific research questions. Figueroa et al. (2023) developed a custom framework combining word embeddings with human coding to create a racism narrative typology, demonstrating how CGT can be adapted to address sensitive healthcare topics. Agudamu et al. (2025) modified standard LDA procedures to include Jaccard similarity calculations for validating topic stability, introducing methodological innovations to ensure robustness of findings.

### Healthcare Domains and Applications

The studies spanned diverse healthcare contexts, demonstrating CGT’s versatility in addressing various health-related research questions. In public health and health promotion, three studies focused on population-level health issues. The COVID-19 risk perception study by Adekunle et al. (2025) revealed how online communities develop distinct risk narratives, with important implications for public health messaging. Their analysis showed that communities such as r/LockdownSkepticism and r/Masks4All developed fundamentally different conceptualizations of COVID-19 risk, driven by underlying ideological commitments rather than simply different interpretations of scientific evidence. The #Movember analysis by Berriche et al. (2024) identified four segments of individual commitment to health campaigns, ranging from sympathizers to maintainers, each requiring different engagement strategies.

Digital health and technology adoption emerged as another major application area. Multiple studies examined how users perceive and adopt health technologies. Weber et al. (2024) analyzed medical professionals’ perceptions of AI, identifying themes of job replacement anxiety and knowledge gaps that influence technology acceptance. Their findings revealed a complex relationship between AI knowledge and adoption intentions, moderated by fears about professional displacement. Guo et al. (2024) examined FemTech applications, revealing six primary affordances perceived by users: instrumental, engagement, aesthetics, self-enhancement, community support, and partner support. These findings provide concrete guidance for technology developers seeking to create user-centered health applications.

Health equity and social justice applications demonstrated CGT’s capacity to address critical social issues in healthcare. Figueroa et al. (2023) applied CGT to examine racism narratives in medical literature, developing a framework for classifying narratives along an anti-racism spectrum from dismissal to actionable anti-racism. Their analysis revealed how medical literature perpetuates or challenges racist narratives through subtle linguistic choices and framing devices. This application shows how CGT can contribute to efforts to achieve health equity by making visible the narratives that shape medical knowledge and practice.

Clinical and health system applications illustrated CGT’s relevance to direct patient care and system improvement. Studies addressing clinical contexts included diabetes management (Vidyadharan et al., 2024) and the use of AI assistance in clinical practice (Yue et al., 2025). The diabetes study identified key themes in patient self-management strategies, while the AI assistance study provided practical guidance for integrating computational support into clinical research. Agudamu et al. (2025) demonstrated health system applications through their analysis of China’s old-age social welfare system, identifying seven key policy themes that could guide system reform.

### Methodological Innovations and Approaches

The reviewed studies demonstrated several methodological innovations in applying CGT to healthcare research (Table 3). Most studies employed multi-phase analytical approaches consistent with Nelson’s (2020) CGT framework, typically involving three key stages. Pattern detection using computational techniques to identify initial patterns in the data served as the foundation, followed by pattern refinement through human interpretation to contextualize and validate computational findings. Finally, pattern confirmation through additional computational or qualitative analysis ensured robustness of results. This three-phase approach was adapted across studies to fit specific research contexts and questions.

**Table 3.** Methodological Innovations in CGT Application.

| Innovation | Study | Description | Contribution to CGT |
| --- | --- | --- | --- |
| Three-phase iteration | All studies | Pattern detection → Refinement → Confirmation | Establishes standard workflow |
| Human-AI collaboration | Yue et al. (2025) | Structured integration of ChatGPT | Demonstrates AI augmentation potential |
| Mixed computational methods | Weber et al. (2024) | Clustering + qualitative analysis | Shows complementary approaches |
| Theory-driven coding | Figueroa et al. (2023) | Computational patterns + theoretical framework | Bridges induction and deduction |
| Validation metrics | Agudamu et al. (2025) | Jaccard similarity for topic stability | Ensures reproducibility |
| Multi-level analysis | Adekunle et al. (2025) | Community → Theme → Concept | Captures hierarchical patterns |

A key theme across all studies was the careful integration of human expertise with computational analysis. Rather than viewing computational techniques as replacements for human interpretation, successful CGT applications demonstrated iterative collaboration between algorithmic pattern detection and expert knowledge. Yue et al. (2025) explicitly examined human-AI collaboration in qualitative analysis, finding that ChatGPT could enhance coding efficiency while requiring human oversight for depth and context. Their study provided detailed guidance on how researchers can effectively collaborate with AI systems, including strategies for prompt engineering, result validation, and maintaining interpretive control.

Studies leveraged diverse digital data sources previously inaccessible to traditional qualitative research, expanding the scope of healthcare research. Reddit discussions featured prominently in three studies, providing access to naturally occurring health-related discourse. Twitter data enabled real-time analysis of health campaign engagement. App reviews offered insights into user experiences with health technologies. Medical literature provided a corpus for analyzing professional discourse about health equity. Interview transcripts demonstrated CGT’s applicability to traditional qualitative data. This diversity of data sources illustrates CGT’s flexibility in addressing different types of healthcare research questions.

Several studies introduced novel analytical techniques that advance CGT methodology. Weber et al. (2024) developed a mixed-method approach combining computational clustering with in-depth qualitative analysis, demonstrating how CGT can incorporate multiple analytical perspectives. Figueroa et al. (2023) created a custom coding framework that integrated computational pattern detection with theory-driven categorization, showing how CGT can be adapted for theory-testing as well as theory-generation. Agudamu et al. (2025) introduced validation techniques using Jaccard similarity to ensure stability of topic models, addressing concerns about reproducibility in computational research.

### Key Findings and Theoretical Contributions

The studies generated significant theoretical insights through CGT application, demonstrating the method’s capacity for both discovery and theory development.

Several studies developed new typologies or frameworks that advance understanding of healthcare phenomena. Adekunle et al. (2025) identified three key themes in COVID-19 risk perception: consequences of AI, physician-AI relationship, and proposed ways forward. Their analysis revealed how risk perceptions are shaped by community identity and ideological commitments rather than simply information processing. Figueroa et al. (2023) created a racism narrative framework with 4 broad categories and 12 granular modalities, providing a comprehensive system for analyzing how medical literature discusses racial health inequities. Guo et al. (2024) identified six FemTech affordances, offering a theoretical framework for understanding user engagement with women’s health technologies.

CGT enabled discovery of patterns not apparent through traditional analysis methods. Studies consistently reported identification of latent themes and relationships that would have been difficult to detect manually. Weber et al. (2024) revealed the moderating role of AI knowledge on job replacement anxiety among medical professionals, a nuanced relationship that emerged through computational analysis of large-scale discourse. Berriche et al. (2024) identified a three-element structure of social movement commitment (segments, emotions, and cognitive factors) that provides a more complex understanding of health campaign engagement than previous models. These discoveries demonstrate CGT’s capacity to reveal hidden structures in healthcare discourse.

Studies also demonstrated how CGT can refine and extend existing theories. Several researchers used CGT findings to elaborate theoretical frameworks from other domains. The application of affordance theory in digital health contexts by Guo et al. (2024) extended understanding of how users perceive and engage with health technologies. The extension of risk perception theories to online communities by Adekunle et al. (2025) showed how traditional theories must be adapted for digital contexts. The development of anti-racism frameworks in medical discourse by Figueroa et al. (2023) provided new theoretical tools for understanding health equity. These theoretical contributions show how CGT can bridge empirical findings with abstract conceptualization.

### Advantages of CGT in Healthcare Research

The reviewed studies consistently identified several advantages of CGT over traditional qualitative methods (Table 4). Scalability emerged as a primary benefit, with CGT enabling analysis of datasets far exceeding traditional qualitative capacity. Studies analyzed hundreds of thousands of documents, with Adekunle et al. (2025) examining 500,000 Reddit comments and Berriche et al. (2024) analyzing 144,906 tweets. This scale of analysis would be impossible with manual coding methods, yet CGT maintained the interpretive depth characteristic of qualitative research. The ability to analyze large datasets is particularly valuable in healthcare research, where understanding population-level patterns while maintaining sensitivity to individual experiences is crucial.

**Table 4.** Advantages and Challenges of CGT in Healthcare Research.

| Aspect | Advantages | Challenges | Mitigation Strategies |
| --- | --- | --- | --- |
| Scale | • Analyze 100,000s of documents<br>• Population-level insights<br>• Comprehensive coverage | • Information overload<br>• Difficulty prioritizing findings | • Hierarchical analysis<br>• Human-guided filtering |
| Pattern Detection | • Discover latent themes<br>• Identify subtle relationships<br>• Temporal trend analysis | • May miss context<br>• Risk of spurious patterns | • Iterative validation<br>• Domain expert review |
| Reproducibility | • Transparent procedures<br>• Replicable analysis | • Parameter sensitivity<br>• Platform dependencies | • Detailed documentation |
|  | • Cumulative knowledge |  | • Open-source tools |
| **Theory Development** | • Novel frameworks<br>• Cross-domain insights<br>• Hypothesis generation | • Theory-data gap<br>• Over-interpretation risk | • Strong theoretical grounding<br>• Multiple analyst validation |
| **Resources** | • Efficiency at scale<br>• Automated processing | • Technical expertise needed<br>• Computational infrastructure | • Training programs<br>• Collaborative teams |

Pattern discovery capabilities of CGT revealed insights that human coders might miss. Computational techniques excel at identifying subtle patterns across large datasets, including latent topics in unstructured text that may not be immediately apparent to human readers, subtle linguistic patterns indicating bias or assumptions in medical discourse, and temporal trends in health-related discussions that emerge over extended time periods. These pattern discovery capabilities are particularly valuable for identifying emerging health concerns, tracking the evolution of health beliefs, and understanding how health information spreads through communities.

Enhanced reproducibility compared to purely interpretive methods addresses a key limitation of traditional qualitative research. Computational components of CGT can be documented and replicated, enabling other researchers to verify findings or apply similar methods to new datasets. Studies provided detailed documentation of computational procedures, including algorithm selection, parameter settings, and validation methods. This transparency enhances the credibility of qualitative research and enables cumulative knowledge building across studies.

The combination of computational pattern detection and human interpretation generated novel theoretical insights across all studies. CGT’s ability to process large amounts of data while maintaining theoretical sensitivity enabled researchers to identify patterns that inform theory development. Studies reported developing new conceptual frameworks, identifying previously unrecognized relationships between concepts, and generating hypotheses for future research. This theoretical innovation demonstrates CGT’s value not just for description but for advancing conceptual understanding of healthcare phenomena.

### Challenges and Limitations

Despite these advantages, studies identified several challenges in implementing CGT. Technical complexity emerged as a significant barrier, with successful CGT implementation requiring expertise in both computational methods and qualitative research. Researchers needed understanding of machine learning algorithms, programming skills for data processing and analysis, and ability to interpret and validate computational outputs. This technical requirement may limit accessibility for researchers without computational backgrounds, potentially creating disparities in who can conduct CGT research.

Balancing computational results with meaningful interpretation remains an ongoing challenge. Several studies noted difficulties in ensuring that computational patterns translate to theoretically meaningful insights. The risk of being overwhelmed by computational outputs without adequate theoretical grounding was acknowledged across studies. Researchers emphasized the need for strong theoretical frameworks to guide interpretation of computational findings, highlighting that CGT is not simply about applying algorithms but about meaningful integration of computational and interpretive approaches.

Context loss represents another limitation, with some studies noting that computational techniques might miss contextual nuances captured by traditional qualitative methods. While computers excel at pattern detection across large datasets, they may miss subtle contextual factors that influence meaning, implicit cultural assumptions that shape discourse, and non-textual elements of communication such as tone or emphasis. Researchers addressed this limitation through careful human interpretation, but acknowledged that some contextual richness may be lost in computational processing.

Resource requirements for CGT implementation can be substantial. Beyond technical expertise, CGT often requires significant computational resources for processing large datasets, interdisciplinary collaboration between technical and domain experts, and time for iterative refinement between computational and interpretive phases. These resource requirements may limit CGT adoption in resource-constrained settings or for time-sensitive research questions.

## Discussion

This systematic review reveals that Computational Grounded Theory (CGT) represents a significant methodological advancement in healthcare research, successfully bridging computational analysis with interpretive depth. The eight reviewed studies demonstrate CGT’s versatility across diverse healthcare domains—from COVID-19 risk perception to health technology adoption—indicating its broad applicability for analyzing complex health phenomena.

The predominance of Latent Dirichlet Allocation (LDA) across studies (6/8) suggests its utility as a foundational technique for pattern detection in healthcare texts. However, the successful integration of diverse computational methods—including sentiment analysis, word embeddings, and deep learning—demonstrates that technique selection should align with specific research questions. The evolution toward more sophisticated approaches, as seen in the AI perception studies, indicates a maturing methodological landscape.

Critical to CGT’s success is the iterative integration of human expertise with computational analysis. Rather than replacing human interpretation, the most effective applications employed cycles where computational techniques surfaced patterns for human interpretation, while domain expertise guided computational refinement. This synergy addresses longstanding tensions between qualitative depth and quantitative breadth in healthcare research.

The studies generated significant theoretical contributions, developing new frameworks for understanding technology affordances, racism narratives, and risk perception in digital contexts. CGT’s capacity to analyze large-scale data while maintaining theoretical sensitivity enabled insights impossible through either computational or qualitative methods alone. Practical applications emerged across intervention design, technology development, and policy formulation, demonstrating CGT’s value beyond academic inquiry.

However, important limitations persist. The relatively small number of included studies reflects the nascent state of CGT in healthcare but limits generalizability of findings. Additionally, the quality assessment criteria had to be adapted for this novel methodology, as standard qualitative or quantitative assessment tools were not fully applicable. Technical complexity remains a barrier, potentially limiting adoption among healthcare researchers lacking computational training. Balancing computational outputs with meaningful interpretation requires strong theoretical grounding. Context loss in computational processing must be weighed against scalability benefits. Resource requirements for infrastructure and interdisciplinary collaboration may restrict accessibility in resource-constrained settings.

## Conclusions

CGT represents a transformative methodological innovation for healthcare research, offering unique capabilities for analyzing large-scale textual data while maintaining theoretical depth. This review demonstrates successful applications across diverse healthcare domains, with studies consistently reporting enhanced pattern discovery, theoretical innovation, and practical insights compared to traditional methods.

Key strengths include scalability to datasets exceeding traditional qualitative capacity, discovery of latent patterns through computational analysis, enhanced reproducibility through transparent procedures, and capacity for theoretical development through human-computer collaboration. CGT proves particularly valuable for understanding rapidly evolving health contexts, analyzing diverse stakeholder perspectives, and generating actionable insights for healthcare improvement.

Future development should focus on standardizing methodological procedures, developing user-friendly tools to lower technical barriers, expanding applications to emerging healthcare challenges, and ensuring equitable access across research settings. Addressing current limitations requires investment in training programs, development of healthcare-specific computational tools, and establishment of ethical frameworks for large-scale health data analysis.

As healthcare systems generate ever-increasing volumes of textual data, CGT provides an essential methodological framework for transforming this information into meaningful insights. By thoughtfully integrating computational power with interpretive wisdom, CGT offers a path forward for healthcare research that honors both the complexity of human health experiences and the opportunities presented by digital data. This systematic review provides a foundation for future CGT applications while highlighting the need for continued methodological development to realize its full potential in advancing human health and wellbeing.

## Data Availability

All data produced in the present work are contained in the manuscript

## Notes

### Competing Interest Statement

The authors have declared no competing interest.

### Funding Statement

This study did not receive any funding

